# How social engagement against Covid-19 in a Brazilian Slum helped mitigate rising statistics

**DOI:** 10.1101/2021.01.06.21249243

**Authors:** Heitor Evangelista, Sérgio J. Gonçalves, Eduardo Delfino Sodré, Juliana Nogueira, Rodrigo Goldenberg-Barbosa, Newton Magalhães, Angela M.G. dos Santos, Ricardo H.M. Godoi, Cesar Amaral, Marcio Cataldo G. da Silva, Daniel A. Junger de Oliveira, Luís Cristóvão Porto

## Abstract

**Objectives:** For many underdeveloped countries, strategies implemented by social communities allied to scientific knowledge may be a rote to attenuate the rapid spread of Covid-19 cases and allow services to the population. This work presents a joint effort collaboration between scientists and underserved community groups from a Brazilian slum/Santa Marta in Rio de Janeiro City in the fight against Covid-19. Measurements of contamination in the air near the ground, georeferencing of data of infected people, were regressed with sanitization activities aiming at reducing the Covid-19 incidence.

**Methods:** We monitored aerosol containing SARS-Cov-2 virus in outdoor ambient air using various virus collection mediums (solid, liquid, and gelatinous substrates) at different aerodynamic sizes. We implemented a local statistics survey for the Covid-19 database correlated with varying sanitization levels between April 2020 and June 2021 developed in the Santa Marta slum.

**Findings:** We detected the SARS-CoV-2 virus in the air near the ground in diameters ranging from 0.25 to 0.5 µm, demonstrating that there is a circulation of the virus in the slum atmosphere. We demonstrate that Covid-19 cases for the Santa Marta slum were significatively lowered with improved sanitization levels (r = −0.74).

**Conclusions:** Despite previous publications that discarded the use of sanitization as a relevant tool in the fight against Covid-19, our results suggest that profits can be achieved in mitigating Covid-19 in underserved community sites. Furthermore, a permanent sanitization activity may induce positive social behavior for the sake of combating Covid-19.

## 1. Introduction

In Brazil, around 13.6 million people, which represents 6% of the population, live in slums (“favelas”) or similar agglomeration households, according to the latest official 2019 census by the Brazilian Institute of Geography and Statistics (IBGE). More than 13 thousand slums were identified throughout Brazil in 734 of the 5,570 Brazilian municipalities. According to the World Health Organization (WHO), 1 billion people live in such conditions around the globe. It is estimated that nearly 89% of people living in slums are concentrated in capital cities and metropolitan regions. In these places, community health is impacted by factors arising from narrow shared physical and social spaces despite a lack of continued social health care. In Brazil, and predominantly in the City of Rio de Janeiro, slums were historically established and lately enlarged on the slopes of hills, following the complex geomorphology of the city landscapes.

Since the begging of the Covid-19 outburst, several studies pointed that different contamination risks exist when considering air quality of indoor and outdoor places(1–3). For the SARS-CoV-2 virus, different proposed mechanisms of transmissibility have been discussed to explain the relationship between the aerial contamination virus on air and the Covid-19 incidence(2,4,5). In this context, the transmission of the pathogens by aerosols or droplets has been shown to be highly predominant in confined spaces(6). The WHO and the US Centers for Disease Control and Prevention (7) have officially recognized inhaling virus-laden aerosols as the primary mode of transmission of COVID-19. Depending on the stage of the epidemiological curve and when hospitalizations reached controlled levels, some countries adopted temporary outdoor maskless conditions for their citizens despite the risks involved.

Due to its architecture of pilled habitations, the outdoor environment of slums in Rio de Janeiro has poor ventilation, high humidity and is mainly shaded by walls which gives them characteristics closer to indoor-type conditions. In addition to that, we find open sky sewage ditches in between narrow alleys. The sewage discharged on the inclined plane of the slums produces water flow turbulence resulting in water splashes. In this condition, sewage droplets microparticles are launched and dispersed in the slum’s atmosphere, reaching the inside of homes (where people do not use masks) and encompassing the walkways passersby use daily, where kids play and do their social life. Several countries of Europe, the USA, Australia, and Brazil(8–12) have reported detecting the SARS-CoV-2 virus in sewage samples. Diarrhea has been described as a symptom of Covid-19 disease, and human excreta may contain important viral loads derived by infected people(13).

The stability of infectious SARS-CoV-2 to a variety of porous, non-porous surfaces and suspended in suspension in the air near the ground have been put into perspective, highlighting this information on the need for hygiene. According to Chin et al. (2020)(14), the SARS-CoV-2 is susceptible to many standard disinfection methods. However, sanitization efficiency against living microorganisms in the environment depends on several factors, some intrinsic properties of the microorganism, others depending on the external physical characteristics of the environment. Only surfaces in direct contact with the disinfection product are predicted quantities will be successfully sanitized. It is believed that the same disinfectants can kill different SARS-CoV-2 variants because no genetic changes to the virus were observed concerning their basic outer layer (envelope) of proteins and lipids(15).

Experiments performed under controlled laboratory conditions regarding the environmental survival of SARS-CoV-2 on surfaces do not necessarily reflect real-world situations since the initial virus load, and factors related to its removal or degradation are frequently underestimated(16). The present work aims to present a case study of sanitization effectiveness, developed in a slum area of Rio de Janeiro City/Santa Marta slum, between April 2020 and June 2021, that considered the 3 main parameters: the presence/absence of SARS-CoV-2 particles in the outdoor ambient, the statistics of Covid-19 cases in studies location and the different levels of sanitization activities.

## 2. Methods

### 2.1 Covid-19 epidemiological database

The Covid-19 epidemiological database is centralized by the “Painel Rio COVID-19” governmental program. Data is obtained in daily resolution for the State of Rio de Janeiro, Brazil, and available at the site “https://experience.arcgis.com/experience/38efc69787a346959c931568bd9e2cc4/”. A second epidemiological program, named “Covid por CEP/Rio de Janeiro,” uses the ZIP-Code of each infected individual and georeferenced its location over a map of the municipality. From this approach, we may select the epidemiological data exclusively from the Santa Marta slum site (Fig. 1). Santa Marta slum comprises a population of approximately 7,356 habs with a density of 134,000 habs km^-2^ in the Southern sector of the City of Rio de Janeiro, Brazil. Our study comprises the period from April 14^th^, 2020, to June 1^st^, 2021. A survey developed by the NOIS program (Núcleo de Operações e Inteligência em Saúde) estimated that Covid-19 notification in Rio de Janeiro is only 7.2%.

**Figure 1.**
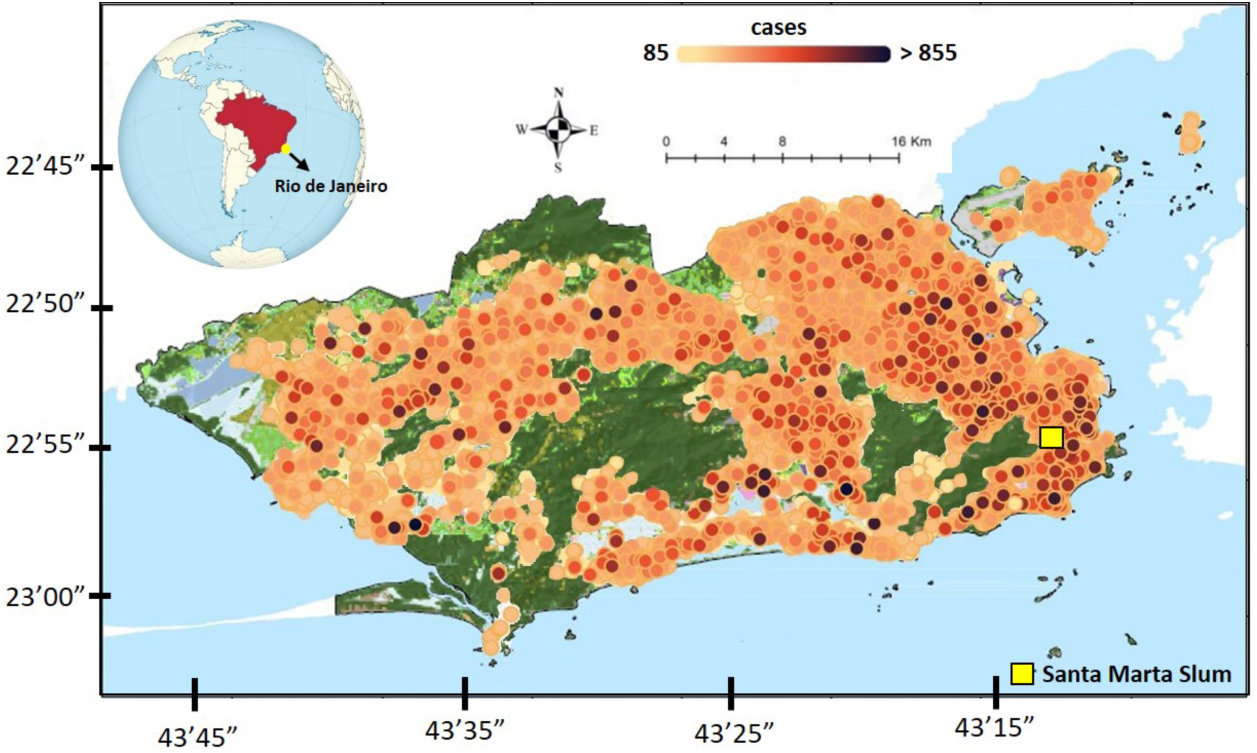
Map distribution of Covid-19 incidence in the City of Rio de Janeiro-Brazil. Data is updated daily and available online by the City Hall/SIVEP-Gripe at https://covidporcep.rio.br/. Green areas denote natural vegetation. The small yellow square indicates the location of the Santa Marta slum.

### 2.2 Sanitization activities

At Santa Marta slum, a team of local volunteers conducted the sanitization activities, dressing appropriate clothes and masks and using cationic surfactants based on quaternary ammonium compounds (5^th^ generation), (QACs), and sodium hypochlorite. Disinfectant spraying occurred manually at floors and surfaces of walls and corridors. The quaternary ammonium belongs to a family of strong antimicrobial compounds and has powerful disinfectant activity, acting by destroying the microorganism’s proteins. It acts directly on the plasma membrane or bacterial cell wall, inhibiting its synthesis. In this work, we have categorized the sanitization levels according to the number of monthly activities. 8 sanitization activities corresponded to 100% (2 per week), 4 to 50%, 3 to 37.5% and 1 to 12.5%. Total sanitization was divided into six campaigns: April (day 14)-August 2020; September-November 2020; December 2020; January-March 2021; April 2021, and May 2021.

### 2.2. Aerosol sampling

For the air samplings, we have used a set of field instrumentation appropriate to virus collection that employed solid, liquid, and gelatinous substrates taking into account previous protocols conducted at the City of Wuhan in China and other sites by the end of 2019 (4,17). Samplings were conducted between July and September 2020, integrating approximately 24 hours between sanitization activities. Besides the Santa Marta slum, a second slum named Rocinha (population of 100,000 habs and density: 70,000 habs km^-2^) was also monitored for comparison purposes. A summary of the methods is presented below.

#### 2.2.1. One Stage Bioaerosol Impactor

The Model N6 (also known as “Andersen head”) is instrumentation that allowed continuous collection of viable bioaerosols and microorganisms in general in the air under a controlled air flux of 12.5 L min^-1^ (18). Bioaerosols were collected in sizes ranging from 0.6 to 22 µm and deposited into a standard Petri dish containing agar.

#### 2.2.2. “BioSampler” (or bioflask)

This sampler consists of a flask subdivided into three compartments: collector spout, a specially designed microorganism trap located in its central part, and an air outlet (19). The collector nozzle is a moving part that functions as an “inlet” through which atmospheric air passes. This compartment is fitted over a central part containing tangential nozzles designed to form a vortex inside the sampler. A liquid solution (glycerol) was filled this inner compartment where bioaerosols could be trapped.

#### 2.2.3. MAS100 sampler

This sampler uses the same principle as the Andersen sampler. It is a compact and integrated suction system in which air is forced through a flat cover with 400 equally spaced holes, which acts as an “inlet” (20). The diameter of each hole is 0.7 mm. Just below the holes, a Petri dish containing a solid culture medium (agar) was arranged so that the bioaerosols from the suction of the air directly impact the Petri dish surface containing agar.

#### 2.2.4. Cascade Impactor

A selective instrumentation for airborne particles that collects particles according to their aerodynamic diameter under a constant airflow of 1.5 L min^-1^. In this work, we used a PIXE impactor that allowed discriminate bioaerosols according to their aerodynamic size as follows: 16µm to 8µm; 8µm to 4µm; 4µm to 2µm; 2µm to 1µm; 1µm to 0,5µm; 0,5µm to 0,25µm; 0,25µm to 0,12µm; 0,12µm to 0,06µm and < 0,06µm. Bioaerosols were deposited on PTFE filters in 24 hours of integration.

### 2.3. Molecular analysis

The samples collected on PTFE filters, agar, and glycerol were transported in cryotubes and refrigerated to laboratory conditions. 2 mL of saline solution was added to each cryotube and left for 30 min for absorption. After that period, the samples were mixed by vortexing for 1 min to prepare the extraction. The material was extracted with the commercial Bio gene DNA/RNA viral (Bioclin®) to extract viral nucleic acid by the silica membrane method. Extracts were eluted in 70 μL of each sample as recommended by the kit. 5µl of the previously extracted RNA was removed and added to 10µl of the reaction mixture containing the target/primer E (viral envelope) and Rp (human control) of the SARS-Cov-2 molecular kit (Bio-Manguinhos), according to the manufacturer’s recommendations. Reverse transcription and amplification reactions and detection of the Coronavirus viral RNA were performed in an Applied Biosystems™ 7500 Real-Time PCR System or QuantStudio™ 5 Real-Time PCR System thermocycler (Applied Biosystems, USA) and programmed for 50°C/15min; 95°C/2min, 95°C/20s, 58°C/30s (45 cycles). According to the Berlin protocol, configure the qRT-PCR parameters with the SARS-Cov-2 molecular kit for SARS-Cov2 (E) - Bio-Manguinhos. RT-PCR was performed with primers and probes directed to the E molecular kit (Bio-Manguinhos), the Threshold cycle (Ct) was adjusted to a fixed limit of 0.2 (E gene) and 0.15 (human control RP) and baseline Start/End range 3–15. The result was considered valid only when the reference gene E’s cycle threshold (Ct) value was ≤ 40.

## 3. Results

For the two campaigns at Santa Marta and Rocinha slum’s, SARS-CoV-2 was detected in the aerosol fraction between 0.25 and 0.5 µm (low virus load – Ct: 40) in the cascade impactor (Fig. 2), where aerosols were discriminated concerning their aerodynamic sizes ranging from 16 µm to smaller than 0.06 µm. The virus detection occurred in a site under sewage spray influence in a corner that forked several alleys. During the monitoring period, Covid-19 epidemiological data had the lowest levels in the entire city of Rio de Janeiro as well as in the Santa Marta slum.

**Figure 2.**
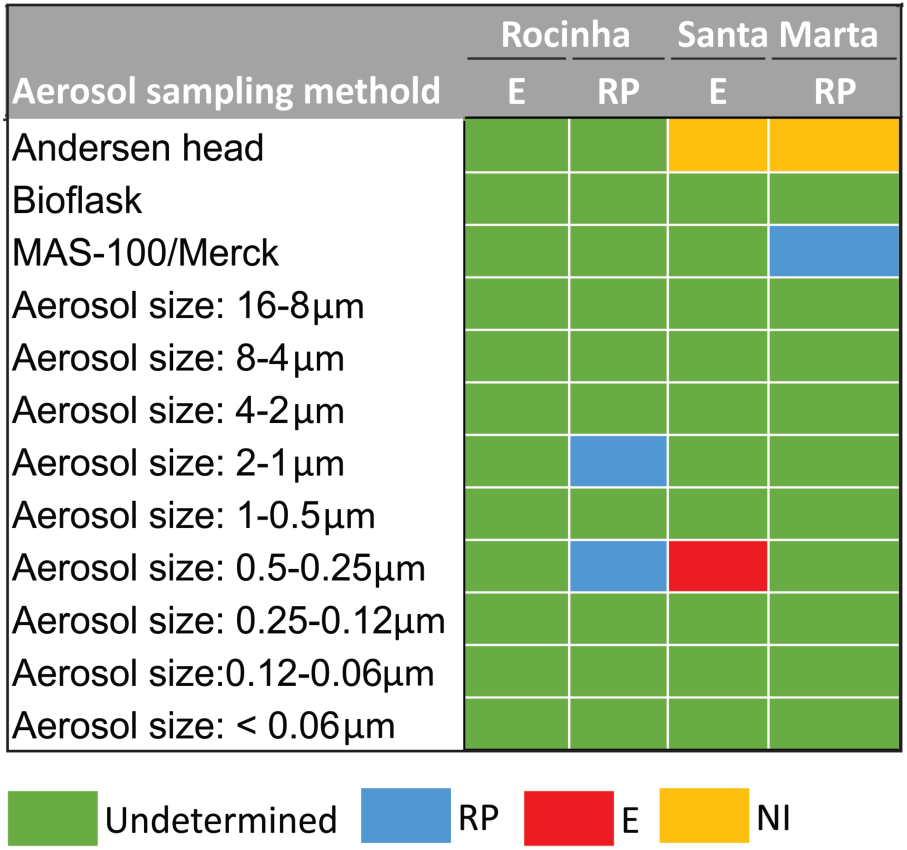
The molecular results for the different types of aerosol sampling techniques used in this work: Andersen head, biosampler, MAS, and a cascade impactor using PTFE 25 mm filters. Colors blue, red, and yellow indicate detection. RP: Gene RdRP, E: Gene E, and NI: Not Inferred.

Considering that the spherical diameter of SARS-CoV-2 ranges from 0.09 to 0.012 µm and its detectability has occurred within the aerosol accumulation mode, we may infer that ambient microparticles infected with the virus have passed through condensation and coagulation processes in the atmosphere before being collected. In droplet transmissions, coughs, sneezes, and speeches can spread course viral particles on saliva and mucus in diameters of approximately 5.0 µm that is much higher than the cutoff size of 0.5 µm particles detected in the Santa Marta slum. Viruses emitted during coughs, sneezes, and speeches are not “particles alone.” They are expelled from the upper respiratory tract in salivary and mucous droplets containing water, salt, protein, and other components of respiratory secretions (21). Their overall sizes may reach an order of magnitude higher than the virus diameter (22). Therefore, the “aerosol component” in the slum atmosphere, in the fine mode, can better explain the viral load detected, which could represent a viral aggregate particle or a small water droplet containing viral particles. In tropical climate conditions, it is expected that these droplets may rapidly dehydrate before they fall down or be attached to other particles in the air. Droplets smaller than 5-10 µm, emitted by human and sewage discharge, tend to dehydrate as fast enough to form droplet nuclei that range within the agglutination airborne size category. These nuclei are easily transported through air currents. In this fraction, virus particles may reach the interior of homes (where people do not wear masks) in the breathable fraction and therefore be able to be deposited as deeper as the bronchiolar lung compartment, according to several lung deposition models (23).

Although it is difficult to precisely whether the viral particles come directed from infected humans or by the spray of sewage, it is the first demonstration that it can survive in the environmental conditions of the slum. From this point, we investigated how effective sanitization could act in the fight against the spread out of the virus. We based this second study on comparing the Covid-19 incidence in the Santa Marta slum and the sanitization level developed in that location.

As illustrated in Figure 3, between April 2020 and June 2021, sanitization in the Santa Marta slum passed through different levels of communitarian support. Once the employed materials depended on public donations and volunteering, the sanitization activity varied from weekly performance (100%) to a complete lack of activity (0%). It covered four SARS-CoV-2 variants in transit in the country. During the first wave of cases and field activities, the original SARS-Cov-2 was already circulating in Rio de Janeiro in April 2020. In September 2020, the Gamma variant (variant P.1) was detected in Manaus/Amazon, and in October, the alpha variant (B.1.1.7) was detected in Brazil. The delta variant (B.1.617.2) came just after, probably arriving in Rio de Janeiro by November or December 2020, approximately one to two months after being detected in India in October 2020. The Beta variant (B.1.351) was identified in December 2020 in South Africa and detected in Brazil in April 2021. Recently, the Omicron variant (variant B.1.1.529) was first reported to WHO(24) on November 2021. Figure 3 presents the Omicron pre-condition period in Brazil.

**Figure 3.**
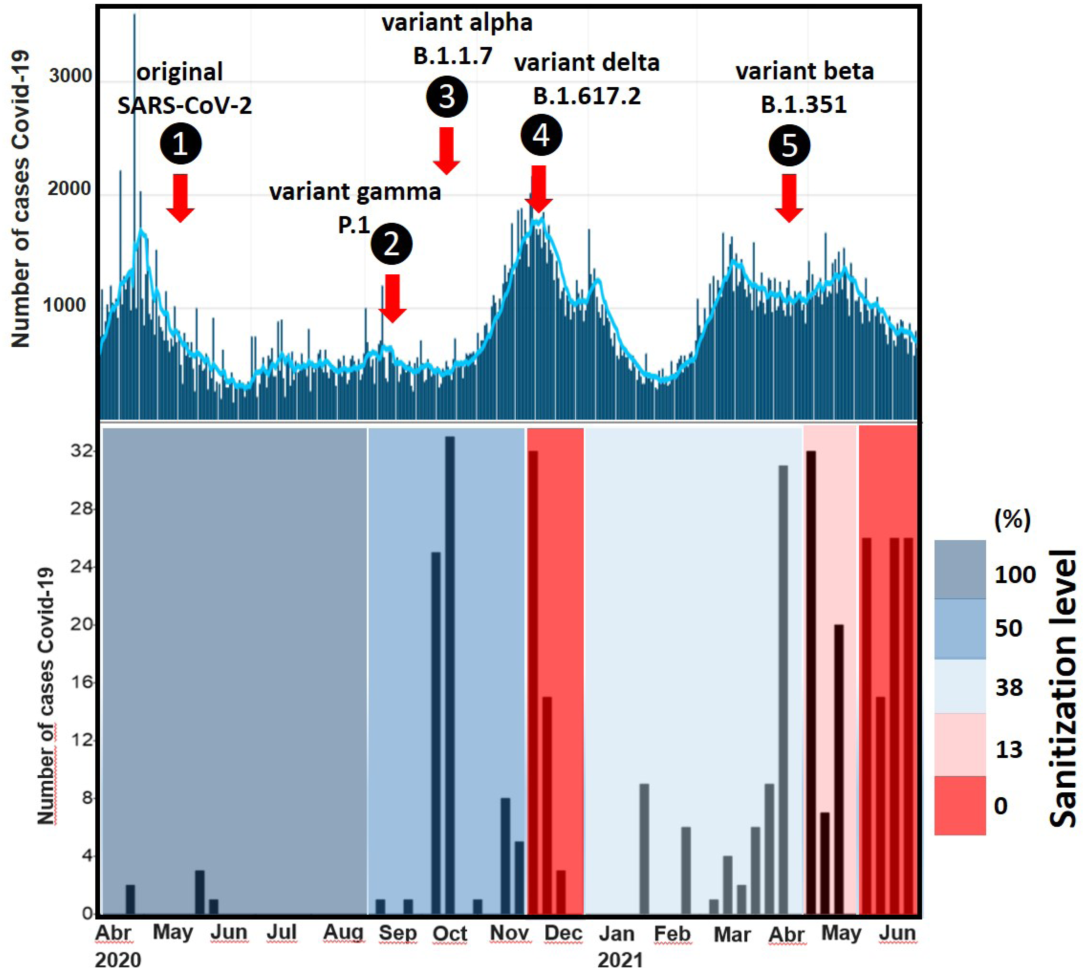
Main SARS-CoV-2 variants under circulation in Rio de Janeiro during the sanitization activities. In the upper box, a 7-day running average of cases in the City of Rio de Janeiro is provided by the Panel COVID-19 (official bulletin from the municipality of Rio de Janeiro (https://riocontraocorona.rio/). Bellow, the time series of accumulated cases in the area of the Santa Marta slum. Shaded boxes correspond to sanitization level. Shaded boxes correspond to sanitization level.

During the first Covid-19 wave in Rio de Janeiro City, from April to August 2020, sanitization activities took place 2 times a week with great social engagement (100% of activity). Within this period, two SARS-CoV-2 variants were detected in Rio de Janeiro. However, despite this fact, the Covid-19 incidence was greatly lower than observed in other sites of the city and other neighboring municipalities. Between September and November, activities halved (50% of activity), and during December, a combination of fatigue, relaxation of social distances, social meetings before and during Christmas, and reduced, or lack, of investments in the project lead to a breakdown in the activities (0% of activity). More than two variants were circulating in the city during December, with a probable prevalence of variant B.1.1.7 (25), which peaked in Brazil in mid-December. The preliminary statistical analysis estimated that this variant is 50% to 74% more transmissible than the original SARS-CoV-2. From January to March 2021, 37.5% (three sanitization activities in a month), relative to the initial level of sanitization, the incidence of Covid-19 was relatively lower during January and February, following the city’s epidemiological curve (upper part of Fig. 3).

Nevertheless, rapid increases were observed when the incidences were higher. In this period, increases in the Santa Marta slum were lagged one month in advance regarding the city database. In April, the same level of sanitization that was carried out since the beginning of January was maintained and likewise the 50% level, and it was not enough to achieve a low level of maintaining the incidence in a low-level pattern. In May and June 2021, sanitization reached its lowest levels of 12.5 and 0%, respectively, and coincidentally, the highest Covid-19 incidence was observed along with the time series (Fig. 3).

There are two points raised from our data of Figure 3: (1) the worst scenario, corresponding to the elevation of cases, that was observed when we experienced 0% sanitization in the same period of, concomitant to the maximum stages of the epidemiological curve that occurred in December 2020 and June 2021; (2) we observed a peak in cases of Covid-19 every time a new variant was documented in the city, regardless of the level of sanitization. We did not find a reason for this response which points to the need for an indeed study concerning other environmental or social factors involved.

## 4. Discussions

Despite the several factors that modulate the incidence of Covid-19, varying from simple use of a mask to complex social behavior, data of Santa Marta slum was significantly correlated with sanitization activities (r = −0.72) (Fig. 4), considering the hygiene work as a whole. In general, the poor areas of the slums in Rio de Janeiro City concentrate some of the highest incidences of Covid-19. A partial socio-epidemiological bulletin for Covid-19 specific for slums (01/2020), edited by the Oswaldo Cruz Foundation/FIOCRUZ (an institution from the Brazilian Ministry of Health), showed that Covid-19 lethality during the first wave in non-slum neighborhoods was estimated to be 9,23% until June 2020, while the neighborhoods areas with high slum concentrations this value climbed to 16,43 to 19,47%. Santa Marta slum was a singular case along with the first wave of cases during the half-year of 2021 when the sanitization effort was maintained at 100% level (2 activities per week). However, in a close look at Figure 4, one may observe that results from the successive sanitization levels of 0, 12.5, 37.5, and 50% did not differ statistically from each other, as seen from the dotted arrows in Figure 4. From this, we may conclude that if sanitization makes any difference on Covid-19 incidence, it should be applied at least two times a week, corresponding to our 100% level. This periodicity conforms to the official recommendations of The Brazilian Ministry of Health for sanitization, using the cationic surfactants ammonium quaternary compound. They suggest a sanitization action two times a week for non-hospitalization sites but of great public circulation as supermarkets, townhouse complexes, and other similar places. Therefore, we believe that the success obtained for the 100% level using QACs is probably related to the extensive use of the product within the expiry time of the disinfectant compound. Locals report from their own experience that an ongoing sanitization activity (when sanitization is maintained at the 100% level), individuals living in the slum tend to be more stimulated to fight against Covid-19 since they have the perception that some activity has been conducted for the sake of collective well-being.

**Figure 4.**
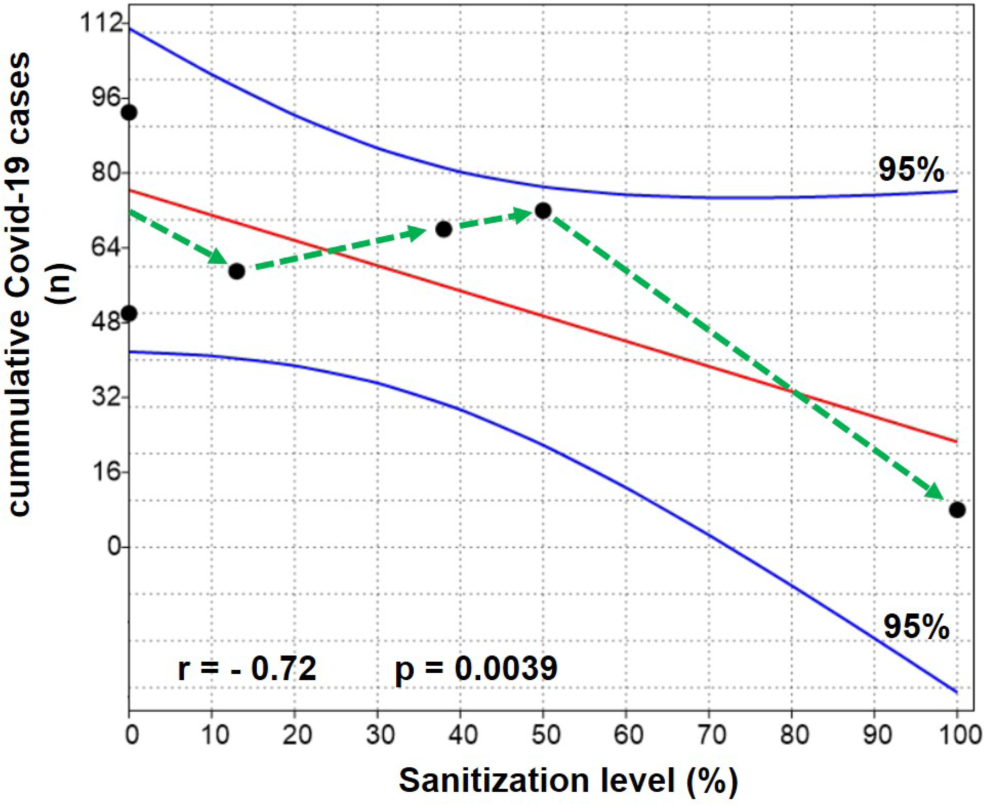
Linear regression (central curve) between the cumulative number of cases of Covid-19 in Santa Marta Slum and the sanitization activities. External curves represent the confidence level of the regression.

One aspect that may favor a good relationship between sanitization level and reduction of cases when considered in a longer time scale (here in 1 year) is that sanitization may bring a better environmental quality condition. Due to a lack of government health care, the slums are home to a series of diseases where can be routinely found a combination of malnutrition, diarrhea, and pneumonia. Diarrhea and pneumonia are conditions often linked to the attack of viruses, bacteria, and fungi. Therefore, an ongoing sanitization may also fight endemic pathogens and thus may provide greater resistance to the immune system of the inhabitants. Our field observation shows that sanitization also contributes to creating a sense of self-preservation and motivation within the communitarians to fight against virus dispersion and contamination.

Another point to be considered in the study is that sanitization activities were conducted during different integration periods, under different SARS-CoV-2 variant circulation and different epidemiological moments. As commented before, these activities happened this way because voluntary activities depend on donations, and there is fatigue of work among the volunteers. In order to consider these aspects and to minimize the impact of the variables cited above, we have also regretted the sanitization levels with the normalized Covid-19 cases, which is defined as the ratio of the integrated number of cases in Santa Marta slum (n) with the total integrated number of cases in the city of Rio de Janeiro (N) in the same period for a given sanitization activity. Data used in this analysis are presented in Table 1. The complete database of Covid-19 for Rio de Janeiro city between April 2020 to June 2021 is shown in Supplementary Material S1. The result obtained (Fig. 5) confirmed the statistically significant response of the Covid-19 incidence with respect the sanitization (r = −0.74), indicating that for the totality of the hygiene activities, a declining trend in cases did occur.

**Table 1.**
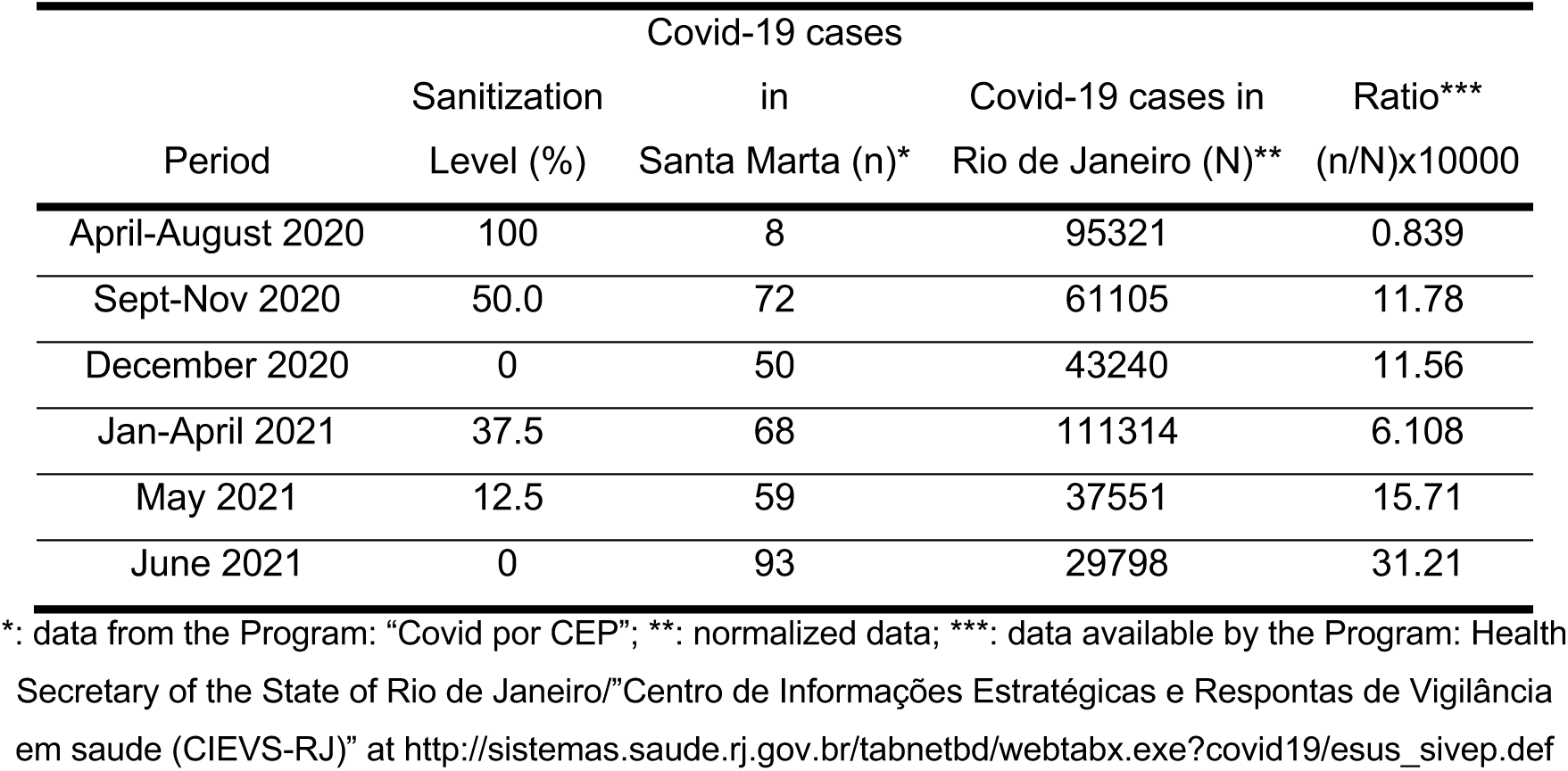
Data of sanitization activities and Covid-19 statistics for Santa Marta slum and the City of Rio de Janeiro.

**Figure 5.**
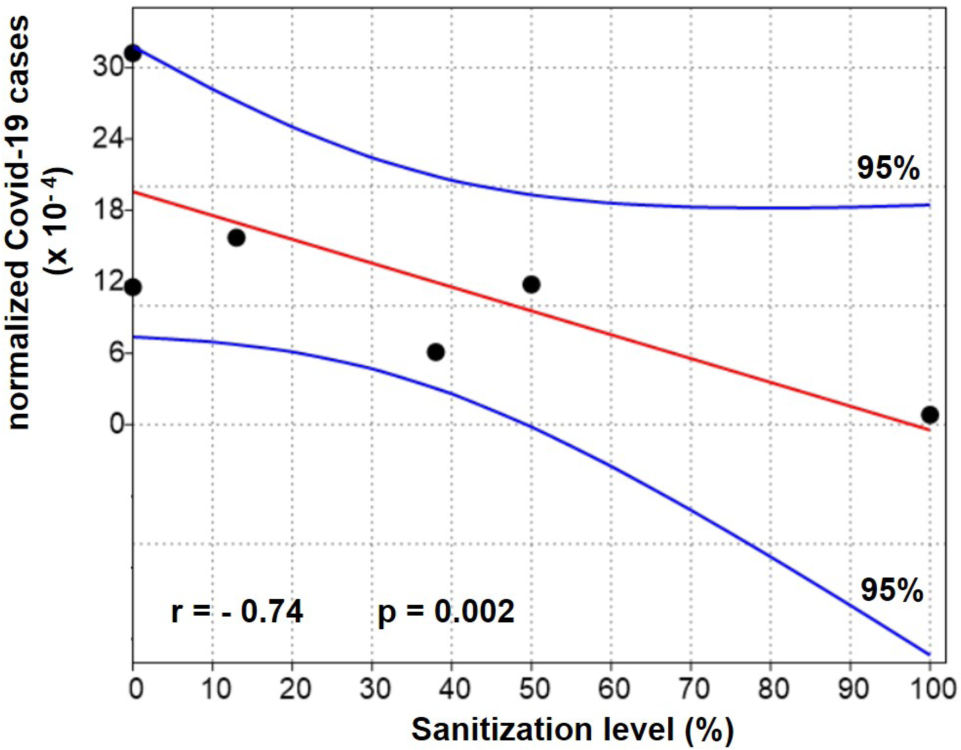
Linear regression (central curve) between the normalized number of cases of Covid-19 (see text) in Santa Marta Slum and the sanitization activities. External curves represent the confidence level of the regression.

Considering the difficulties in maintaining an enduring sanitization program in the Santa Marta slum, which represents an elevated financial cost, a collaboration was established between the local community and the Rio de Janeiro State University-Uerj. By georeferencing Covid-19 individual data, based on a mobile app, and performing aerosol monitoring, the scientific team in the university was able to detect hot spot locations in terms of cases and the ambient air detection of the virus. From this, we constructed risk maps in which the volunteers could focus their sanitization activity to optimize fieldwork performance and use smaller amounts of disinfectant during shorter work schedules. Figure 6a,b shows the spatial distribution of Covid-19 cases in the Santa Marta slum and the resulting product with the application of the kriging method of data interpolation over a satellite image from Google maps of the study site. During the pandemic, Covid-19 cases were more concentrated in the lower parts of the slum where we find common passages of inhabitants and therefore are locations of more intense circulation of people and small markets.

**Figure 6.**
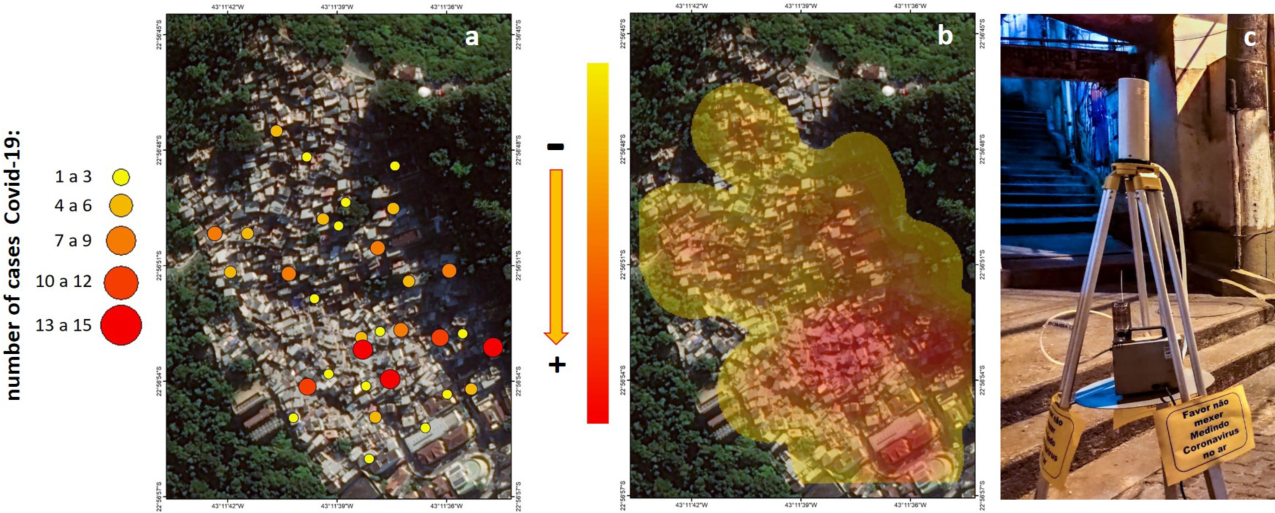
Summary of activities developed at Santa Marta slum: **(a)** spatially distributed of Covid-19 cases in December 21^st^, 2020 (database from https://covidporcep.rio.br/) and georeferencing; **(b)** kriging map and definition of hot spots for sanitization activity; **(c)** aerosol sampling at Santa Marta slum (PIXE sampler).

Owing to mitigate the Covid-19 incidence and optimize the sanitization activity, it is shown in Figure 7 a summary box diagram illustrating the working steps from the data georeferencing to the sanitization optimization in the Santa Marta slum.

**Figure 7.**
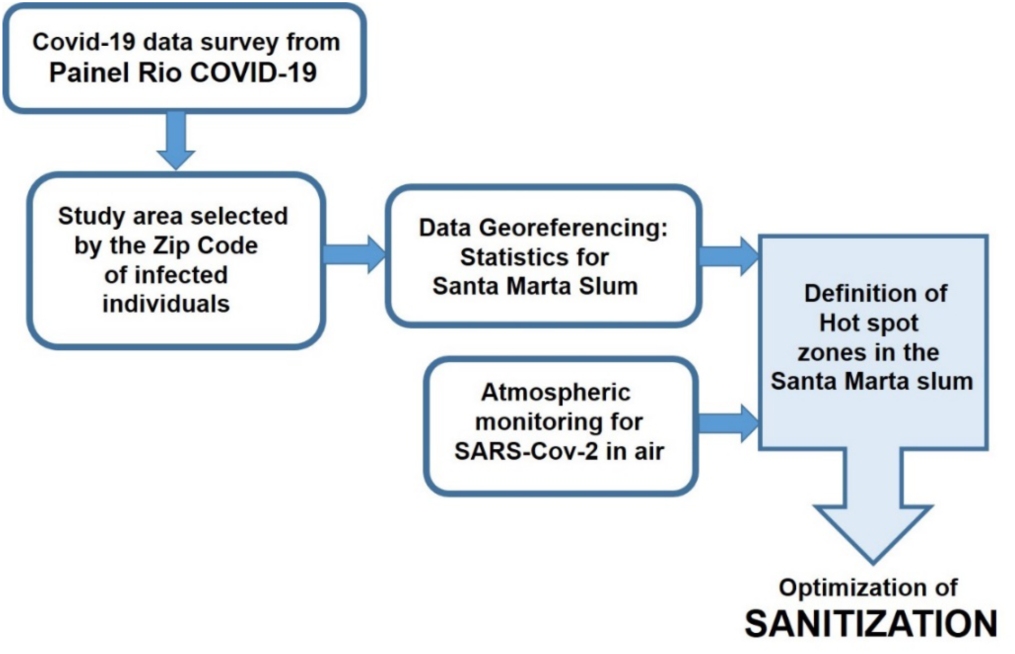
Box diagram illustrating the sequence of activities (database georeferencing and bioaerosols monitoring) for planning and sanitization optimization at Santa Marta slum/Rio de Janeiro City.

## 5. Conclusions

To many underdeveloped countries, Covid-19 highlighted unsolved long-term social-economic problems. Data of *in situ* effectiveness of sanitization activities in outdoor ambient for SARS-CoV-2 is scarce in the present literature and poorly evaluated in real world situations. Although, in general, sanitization has been approached as a controversial activity in terms of effectiveness, our data pointed to real effectiveness in a site of low health care attendance. Our results for Santa Marta slum showed that (1) SARS-CoV-2 was detected in near ground ambient air in diameters ranging from 0.25 to d 0.5 µm demonstrating that there is a circulation of the virus in the slum atmosphere; (2) sanitization activities can provide positive results, especially if it is maintained in high frequency.

## Data Availability

The use of the Santa Marta slum ("favela") database is available for open access at https://covidporcep.rio.br and the Panel Covid-19 (official bulletin from the municipality of Rio de Janeiro) is in https://riocontraocorona.rio.

## Author Contributions

All authors contributed equally to the development of the project and the writing of the manuscript. Specifically, Prof. Heitor Evangelista (Project PI, created the concept of the project and participated in all outdoor samplings and development of methods); Prof. Luís Cristóvão Porto and Ms. Angela M.G. dos Santos performed the rt-PCR analysis; Dr. Sérgio J. Gonçalves Junior, Dr. Juliana Nogueira, MS. Rodrigo G. Barbosa, Dr. Eduardo Delfino Sodré participated and coordinated all outdoor samplings, Dr. Marcio Cataldo Gomes da Silva and Dr. Daniel A.J. de Oliveira added new considerations in the text content and discussions, Dr. Newton Magalhães did the georeferencing methods, Prof. Cesar Amaral revised the text content, discussions and molecular analysis, Prof. Ricardo H.M. Godoi and Dr. Sérgio J. Gonçalves Junior allowed aerosol technology to the project.

## Acknowledgments

We greatly thank Mr. Thiago Firmino and Mr. Thandy Firmino, and the Santa Marta sanitization team for logistics and discussion on routine life and work at slums in Rio de Janeiro City. We thank Ms. Roberta Priori for helping with the map edition and graduate student Enzo H.B.E. da Silva for statistic surveys.

