## Supplementary Material S1 for "How social engagement against Covid-19 in a Brazilian Slum helped mitigate rising statistics"

**PROGRAM "COVID-19 no Rio de Janeiro"**

**Covid-19 Cases in Rio de Janeiro City**

Health Secretaries of the Rio de Janeiro State/Centro de Informações Estratégicas e Respostas de Vigilância em saúde (CIEVS-RJ)

**http://sistemas.saude.rj.gov.br/tabnetbd/webtabx.exe?covid19/esus_sivep.def**

| Day of Notification year/month/day | Rio de Janeiro City CASES (N) |
|---|---|
| **2020/04/01** | 255 |
| **2020/04/02** | 308 |

| | |
|---|---|
| **2020/04/03** | 359 |
| **2020/04/04** | 155 |
| **2020/04/05** | 109 |
| **2020/04/06** | 546 |
| **2020/04/07** | 564 |
| **2020/04/08** | 459 |
| **2020/04/09** | 365 |
| **2020/04/10** | 405 |
| **2020/04/11** | 385 |
| **2020/04/12** | 417 |
| **2020/04/13** | 706 |
| **2020/04/14** | 997 |
| **2020/04/15** | 773 |
| **2020/04/16** | 761 |
| **2020/04/17** | 635 |
| **2020/04/18** | 279 |
| **2020/04/19** | 340 |
| **2020/04/20** | 1,193 |
| **2020/04/21** | 351 |
| **2020/04/22** | 784 |
| **2020/04/23** | 373 |
| **2020/04/24** | 1,282 |
| **2020/04/25** | 391 |
| **2020/04/26** | 311 |
| **2020/04/27** | 1,321 |
| **2020/04/28** | 1,638 |
| **2020/04/29** | 1,063 |
| **2020/04/30** | 1,178 |
| **2020/05/01** | 401 |
| **2020/05/02** | 387 |
| **2020/05/03** | 328 |
| **2020/05/04** | 1,242 |
| **2020/05/05** | 1,131 |
| **2020/05/06** | 1,048 |
| **2020/05/07** | 993 |
| **2020/05/08** | 1,032 |
| **2020/05/09** | 295 |
| **2020/05/10** | 244 |
| **2020/05/11** | 773 |
| **2020/05/12** | 1,080 |
| **2020/05/13** | 1,146 |
| **2020/05/14** | 868 |
| **2020/05/15** | 762 |
| **2020/05/16** | 369 |
| **2020/05/17** | 288 |
| **2020/05/18** | 830 |

| | |
|---|---|
| **2020/05/19** | 1,268 |
| **2020/05/20** | 992 |
| **2020/05/21** | 1,119 |
| **2020/05/22** | 907 |
| **2020/05/23** | 344 |
| **2020/05/24** | 539 |
| **2020/05/25** | 866 |
| **2020/05/26** | 674 |
| **2020/05/27** | 1,191 |
| **2020/05/28** | 981 |
| **2020/05/29** | 876 |
| **2020/05/30** | 391 |
| **2020/05/31** | 472 |
| **2020/06/01** | 946 |
| **2020/06/02** | 822 |
| **2020/06/03** | 981 |
| **2020/06/04** | 725 |
| **2020/06/05** | 582 |
| **2020/06/06** | 186 |
| **2020/06/07** | 203 |
| **2020/06/08** | 835 |
| **2020/06/09** | 867 |
| **2020/06/10** | 793 |
| **2020/06/11** | 232 |
| **2020/06/12** | 789 |
| **2020/06/13** | 297 |
| **2020/06/14** | 338 |
| **2020/06/15** | 1,297 |
| **2020/06/16** | 1,389 |
| **2020/06/17** | 1,585 |
| **2020/06/18** | 983 |
| **2020/06/19** | 1,043 |
| **2020/06/20** | 184 |
| **2020/06/21** | 250 |
| **2020/06/22** | 1,174 |
| **2020/06/23** | 711 |
| **2020/06/24** | 823 |
| **2020/06/25** | 739 |
| **2020/06/26** | 523 |
| **2020/06/27** | 279 |
| **2020/06/28** | 134 |
| **2020/06/29** | 602 |
| **2020/06/30** | 795 |
| **2020/07/01** | 703 |
| **2020/07/02** | 657 |
| **2020/07/03** | 451 |

| | |
|---|---|
| **2020/07/04** | 229 |
| **2020/07/05** | 204 |
| **2020/07/06** | 627 |
| **2020/07/07** | 592 |
| **2020/07/08** | 529 |
| **2020/07/09** | 422 |
| **2020/07/10** | 436 |
| **2020/07/11** | 129 |
| **2020/07/12** | 166 |
| **2020/07/13** | 462 |
| **2020/07/14** | 427 |
| **2020/07/15** | 390 |
| **2020/07/16** | 236 |
| **2020/07/17** | 377 |
| **2020/07/18** | 103 |
| **2020/07/19** | 66 |
| **2020/07/20** | 464 |
| **2020/07/21** | 753 |
| **2020/07/22** | 574 |
| **2020/07/23** | 552 |
| **2020/07/24** | 457 |
| **2020/07/25** | 262 |
| **2020/07/26** | 267 |
| **2020/07/27** | 658 |
| **2020/07/28** | 774 |
| **2020/07/29** | 605 |
| **2020/07/30** | 405 |
| **2020/07/31** | 619 |
| **2020/08/01** | 121 |
| **2020/08/02** | 85 |
| **2020/08/03** | 580 |
| **2020/08/04** | 655 |
| **2020/08/05** | 584 |
| **2020/08/06** | 465 |
| **2020/08/07** | 517 |
| **2020/08/08** | 165 |
| **2020/08/09** | 80 |
| **2020/08/10** | 787 |
| **2020/08/11** | 1,375 |
| **2020/08/12** | 897 |
| **2020/08/13** | 1,364 |
| **2020/08/14** | 754 |
| **2020/08/15** | 184 |
| **2020/08/16** | 168 |
| **2020/08/17** | 906 |
| **2020/08/18** | 1,118 |

| | |
|---|---|
| **2020/08/19** | 943 |
| **2020/08/20** | 808 |
| **2020/08/21** | 804 |
| **2020/08/22** | 145 |
| **2020/08/23** | 397 |
| **2020/08/24** | 791 |
| **2020/08/25** | 804 |
| **2020/08/26** | 594 |
| **2020/08/27** | 483 |
| **2020/08/28** | 457 |
| **2020/08/29** | 179 |
| **2020/08/30** | 94 |
| **2020/08/31** | 774 |
| **2020/09/01** | 748 |
| **2020/09/02** | 699 |
| **2020/09/03** | 614 |
| **2020/09/04** | 484 |
| **2020/09/05** | 275 |
| **2020/09/06** | 157 |
| **2020/09/07** | 137 |
| **2020/09/08** | 801 |
| **2020/09/09** | 755 |
| **2020/09/10** | 1,054 |
| **2020/09/11** | 561 |
| **2020/09/12** | 191 |
| **2020/09/13** | 144 |
| **2020/09/14** | 835 |
| **2020/09/15** | 717 |
| **2020/09/16** | 807 |
| **2020/09/17** | 690 |
| **2020/09/18** | 723 |
| **2020/09/19** | 194 |
| **2020/09/20** | 152 |
| **2020/09/21** | 802 |
| **2020/09/22** | 836 |
| **2020/09/23** | 610 |
| **2020/09/24** | 573 |
| **2020/09/25** | 588 |
| **2020/09/26** | 266 |
| **2020/09/27** | 125 |
| **2020/09/28** | 871 |
| **2020/09/29** | 1,048 |
| **2020/09/30** | 706 |
| **2020/10/01** | 756 |
| **2020/10/02** | 902 |
| **2020/10/03** | 182 |

| | |
|---|---|
| **2020/10/04** | 122 |
| **2020/10/05** | 1,052 |
| **2020/10/06** | 767 |
| **2020/10/07** | 723 |
| **2020/10/08** | 684 |
| **2020/10/09** | 723 |
| **2020/10/10** | 183 |
| **2020/10/11** | 119 |
| **2020/10/12** | 141 |
| **2020/10/13** | 931 |
| **2020/10/14** | 785 |
| **2020/10/15** | 680 |
| **2020/10/16** | 653 |
| **2020/10/17** | 288 |
| **2020/10/18** | 127 |
| **2020/10/19** | 538 |
| **2020/10/20** | 595 |
| **2020/10/21** | 451 |
| **2020/10/22** | 503 |
| **2020/10/23** | 571 |
| **2020/10/24** | 155 |
| **2020/10/25** | 118 |
| **2020/10/26** | 799 |
| **2020/10/27** | 668 |
| **2020/10/28** | 506 |
| **2020/10/29** | 629 |
| **2020/10/30** | 421 |
| **2020/10/31** | 252 |
| **2020/11/01** | 218 |
| **2020/11/02** | 236 |
| **2020/11/03** | 757 |
| **2020/11/04** | 603 |
| **2020/11/05** | 842 |
| **2020/11/06** | 771 |
| **2020/11/07** | 239 |
| **2020/11/08** | 189 |
| **2020/11/09** | 1,063 |
| **2020/11/10** | 976 |
| **2020/11/11** | 814 |
| **2020/11/12** | 880 |
| **2020/11/13** | 948 |
| **2020/11/14** | 325 |
| **2020/11/15** | 204 |
| **2020/11/16** | 1,348 |
| **2020/11/17** | 1,174 |
| **2020/11/18** | 1,153 |

| | |
|---|---|
| **2020/11/19** | 1,183 |
| **2020/11/20** | 457 |
| **2020/11/21** | 443 |
| **2020/11/22** | 283 |
| **2020/11/23** | 1,624 |
| **2020/11/24** | 1,979 |
| **2020/11/25** | 1,519 |
| **2020/11/26** | 1,992 |
| **2020/11/27** | 1,967 |
| **2020/11/28** | 663 |
| **2020/11/29** | 291 |
| **2020/11/30** | 2,777 |
| **2020/12/01** | 2,241 |
| **2020/12/02** | 2,179 |
| **2020/12/03** | 2,008 |
| **2020/12/04** | 1,837 |
| **2020/12/05** | 500 |
| **2020/12/06** | 303 |
| **2020/12/07** | 2,279 |
| **2020/12/08** | 2,400 |
| **2020/12/09** | 2,060 |
| **2020/12/10** | 2,032 |
| **2020/12/11** | 1,800 |
| **2020/12/12** | 331 |
| **2020/12/13** | 277 |
| **2020/12/14** | 1,582 |
| **2020/12/15** | 2,155 |
| **2020/12/16** | 1,717 |
| **2020/12/17** | 1,805 |
| **2020/12/18** | 1,386 |
| **2020/12/19** | 464 |
| **2020/12/20** | 288 |
| **2020/12/21** | 2,031 |
| **2020/12/22** | 1,540 |
| **2020/12/23** | 1,501 |
| **2020/12/24** | 621 |
| **2020/12/25** | 201 |
| **2020/12/26** | 524 |
| **2020/12/27** | 202 |
| **2020/12/28** | 2,565 |
| **2020/12/29** | 2,043 |
| **2020/12/30** | 1,705 |
| **2020/12/31** | 663 |
| **2021/01/01** | 268 |
| **2021/01/02** | 474 |
| **2021/01/03** | 260 |

| | |
|---|---|
| **2021/01/04** | 2,887 |
| **2021/01/05** | 2,160 |
| **2021/01/06** | 1,759 |
| **2021/01/07** | 1,579 |
| **2021/01/08** | 1,420 |
| **2021/01/09** | 456 |
| **2021/01/10** | 251 |
| **2021/01/11** | 1,810 |
| **2021/01/12** | 1,561 |
| **2021/01/13** | 1,294 |
| **2021/01/14** | 1,306 |
| **2021/01/15** | 1,103 |
| **2021/01/16** | 334 |
| **2021/01/17** | 202 |
| **2021/01/18** | 1,519 |
| **2021/01/19** | 1,046 |
| **2021/01/20** | 337 |
| **2021/01/21** | 903 |
| **2021/01/22** | 1,030 |
| **2021/01/23** | 262 |
| **2021/01/24** | 144 |
| **2021/01/25** | 1,187 |
| **2021/01/26** | 963 |
| **2021/01/27** | 917 |
| **2021/01/28** | 806 |
| **2021/01/29** | 620 |
| **2021/01/30** | 186 |
| **2021/01/31** | 249 |
| **2021/02/01** | 758 |
| **2021/02/02** | 718 |
| **2021/02/03** | 581 |
| **2021/02/04** | 577 |
| **2021/02/05** | 568 |
| **2021/02/06** | 185 |
| **2021/02/07** | 141 |
| **2021/02/08** | 754 |
| **2021/02/09** | 603 |
| **2021/02/10** | 640 |
| **2021/02/11** | 579 |
| **2021/02/12** | 525 |
| **2021/02/13** | 143 |
| **2021/02/14** | 85 |
| **2021/02/15** | 545 |
| **2021/02/16** | 190 |
| **2021/02/17** | 746 |
| **2021/02/18** | 673 |

| | |
|---|---|
| **2021/02/19** | 622 |
| **2021/02/20** | 168 |
| **2021/02/21** | 97 |
| **2021/02/22** | 951 |
| **2021/02/23** | 684 |
| **2021/02/24** | 739 |
| **2021/02/25** | 682 |
| **2021/02/26** | 559 |
| **2021/02/27** | 158 |
| **2021/02/28** | 103 |
| **2021/03/01** | 842 |
| **2021/03/02** | 725 |
| **2021/03/03** | 772 |
| **2021/03/04** | 888 |
| **2021/03/05** | 811 |
| **2021/03/06** | 166 |
| **2021/03/07** | 151 |
| **2021/03/08** | 1,365 |
| **2021/03/09** | 1,020 |
| **2021/03/10** | 1,150 |
| **2021/03/11** | 1,051 |
| **2021/03/12** | 1,099 |
| **2021/03/13** | 311 |
| **2021/03/14** | 200 |
| **2021/03/15** | 1,775 |
| **2021/03/16** | 1,385 |
| **2021/03/17** | 1,438 |
| **2021/03/18** | 1,383 |
| **2021/03/19** | 1,328 |
| **2021/03/20** | 468 |
| **2021/03/21** | 286 |
| **2021/03/22** | 2,219 |
| **2021/03/23** | 1,724 |
| **2021/03/24** | 1,466 |
| **2021/03/25** | 1,472 |
| **2021/03/26** | 1,518 |
| **2021/03/27** | 489 |
| **2021/03/28** | 371 |
| **2021/03/29** | 2,092 |
| **2021/03/30** | 1,731 |
| **2021/03/31** | 1,628 |
| **2021/04/01** | 1,474 |
| **2021/04/02** | 593 |
| **2021/04/03** | 539 |
| **2021/04/04** | 301 |
| **2021/04/05** | 2,342 |

| | |
|---|---|
| **2021/04/06** | 1,605 |
| **2021/04/07** | 1,477 |
| **2021/04/08** | 1,484 |
| **2021/04/09** | 1,318 |
| **2021/04/10** | 399 |
| **2021/04/11** | 226 |
| **2021/04/12** | 1,865 |
| **2021/04/13** | 1,326 |
| **2021/04/14** | 1,394 |
| **2021/04/15** | 1,397 |
| **2021/04/16** | 1,450 |
| **2021/04/17** | 426 |
| **2021/04/18** | 277 |
| **2021/04/19** | 1,874 |
| **2021/04/20** | 1,566 |
| **2021/04/21** | 848 |
| **2021/04/22** | 1,408 |
| **2021/04/23** | 952 |
| **2021/04/24** | 392 |
| **2021/04/25** | 240 |
| **2021/04/26** | 2,181 |
| **2021/04/27** | 1,708 |
| **2021/04/28** | 1,417 |
| **2021/04/29** | 1,277 |
| **2021/04/30** | 1,167 |
| **2021/05/01** | 336 |
| **2021/05/02** | 233 |
| **2021/05/03** | 1,779 |
| **2021/05/04** | 1,515 |
| **2021/05/05** | 1,549 |
| **2021/05/06** | 1,576 |
| **2021/05/07** | 1,352 |
| **2021/05/08** | 354 |
| **2021/05/09** | 234 |
| **2021/05/10** | 1,823 |
| **2021/05/11** | 1,630 |
| **2021/05/12** | 1,474 |
| **2021/05/13** | 1,460 |
| **2021/05/14** | 1,402 |
| **2021/05/15** | 573 |
| **2021/05/16** | 284 |
| **2021/05/17** | 2,063 |
| **2021/05/18** | 1,788 |
| **2021/05/19** | 1,653 |
| **2021/05/20** | 1,552 |
| **2021/05/21** | 1,550 |

| | |
|---|---|
| **2021/05/22** | 593 |
| **2021/05/23** | 251 |
| **2021/05/24** | 1,974 |
| **2021/05/25** | 1,610 |
| **2021/05/26** | 1,350 |
| **2021/05/27** | 1,543 |
| **2021/05/28** | 1,247 |
| **2021/05/29** | 582 |
| **2021/05/30** | 306 |
| **2021/05/31** | 1,915 |
| **2021/06/01** | 1,636 |
| **2021/06/02** | 1,535 |
| **2021/06/03** | 417 |
| **2021/06/04** | 1,673 |
| **2021/06/05** | 512 |
| **2021/06/06** | 267 |
| **2021/06/07** | 1,836 |
| **2021/06/08** | 1,551 |
| **2021/06/09** | 1,380 |
| **2021/06/10** | 1,384 |
| **2021/06/11** | 1,303 |
| **2021/06/12** | 340 |
| **2021/06/13** | 169 |
| **2021/06/14** | 1,451 |
| **2021/06/15** | 1,116 |
| **2021/06/16** | 1,228 |
| **2021/06/17** | 1,120 |
| **2021/06/18** | 1,017 |
| **2021/06/19** | 270 |
| **2021/06/20** | 159 |
| **2021/06/21** | 1,611 |
| **2021/06/22** | 1,056 |
| **2021/06/23** | 1,024 |
| **2021/06/24** | 1,179 |
| **2021/06/25** | 1,008 |
| **2021/06/26** | 383 |
| **2021/06/27** | 179 |
| **2021/06/28** | 1,517 |
| **2021/06/29** | 607 |
| **2021/06/30** | 870 |
| **2021/07/01** | 1,021 |
| **2021/07/02** | 914 |
| **2021/07/03** | 243 |
| **2021/07/04** | 171 |
| **2021/07/05** | 1,401 |
| **2021/07/06** | 1,020 |

| | |
|---|---|
| **2021/07/07** | 891 |
| **2021/07/08** | 1,075 |
| **2021/07/09** | 1,036 |
| **2021/07/10** | 328 |
| **2021/07/11** | 214 |
| **2021/07/12** | 1,072 |
| **2021/07/13** | 1,508 |
| **2021/07/14** | 1,146 |
| **2021/07/15** | 1,173 |
| **2021/07/16** | 1,364 |
| **2021/07/17** | 343 |
| **2021/07/18** | 216 |
| **2021/07/19** | 973 |
| **2021/07/20** | 1,601 |
| **2021/07/21** | 1,404 |
| **2021/07/22** | 1,309 |
| **2021/07/23** | 1,319 |
| **2021/07/24** | 439 |
| **2021/07/25** | 234 |
| **2021/07/26** | 2,007 |
| **2021/07/27** | 1,733 |
| **2021/07/28** | 1,618 |
| **2021/07/29** | 1,396 |
| **2021/07/30** | 1,408 |
| **2021/07/31** | 625 |
| **2021/08/01** | 293 |
| **2021/08/02** | 2,181 |
| **2021/08/03** | 1,920 |
| **2021/08/04** | 1,927 |
| **2021/08/05** | 1,972 |
| **2021/08/06** | 1,921 |
| **2021/08/07** | 645 |
| **2021/08/08** | 297 |
| **2021/08/09** | 2,557 |
| **2021/08/10** | 2,637 |
| **2021/08/11** | 2,399 |
| **2021/08/12** | 2,414 |
| **2021/08/13** | 2,093 |
| **2021/08/14** | 779 |
| **2021/08/15** | 319 |
| **2021/08/16** | 3,213 |
| **2021/08/17** | 2,741 |
| **2021/08/18** | 2,543 |
| **2021/08/19** | 2,450 |
| **2021/08/20** | 2,156 |
| **2021/08/21** | 636 |

| | |
|---|---|
| **2021/08/22** | 332 |
| **2021/08/23** | 3,083 |
| **2021/08/24** | 2,362 |
| **2021/08/25** | 2,046 |
| **2021/08/26** | 1,994 |
| **2021/08/27** | 1,934 |
| **2021/08/28** | 578 |
| **2021/08/29** | 244 |
| **2021/08/30** | 2,566 |
| **2021/08/31** | 1,796 |
| **2021/09/01** | 1,594 |
| **2021/09/02** | 1,667 |
| **2021/09/03** | 1,591 |
| **2021/09/04** | 614 |
| **2021/09/05** | 251 |
| **2021/09/06** | 1,640 |
| **2021/09/07** | 283 |
| **2021/09/08** | 1,046 |
| **2021/09/09** | 1,037 |
| **2021/09/10** | 981 |
| **2021/09/11** | 360 |
| **2021/09/12** | 258 |
| **2021/09/13** | 1,467 |
| **2021/09/14** | 1,343 |
| **2021/09/15** | 1,029 |
| **2021/09/16** | 741 |
| **2021/09/17** | 821 |
| **2021/09/18** | 258 |
| **2021/09/19** | 122 |
| **2021/09/20** | 1,219 |
| **2021/09/21** | 679 |
| **2021/09/22** | 752 |
| **2021/09/23** | 664 |
| **2021/09/24** | 642 |
| **2021/09/25** | 208 |
| **2021/09/26** | 118 |
| **2021/09/27** | 869 |
| **2021/09/28** | 569 |
| **2021/09/29** | 485 |
| **2021/09/30** | 541 |
